# SSRI Use During Acute COVID-19 Infection Associated with Lower Risk of Long COVID Among Patients with Depression

**DOI:** 10.1101/2024.02.05.24302352

**Authors:** Zachary Butzin-Dozier, Yunwen Ji, Sarang Deshpande, Eric Hurwitz, Jeremy Coyle, Junming (Seraphina) Shi, Andrew Mertens, Mark J. van der Laan, John M. Colford, Rena C. Patel, Alan E. Hubbard, the National COVID Cohort Collaborative (N3C) Consortium

**Affiliations:** School of Public Health, University of California, Berkeley, Berkeley, CA USA; University of Colorado Anschutz Medical Campus, Aurora, CO USA; University of Alabama at Birmingham, Birmingham, AL USA

**Author notes:** Correspondence: Zachary Butzin-Dozier, PhD, MPH, Division of Biostatistics, School of Public Health, University of California, Berkeley Berkeley Way West, 2121 Berkeley Way, Berkeley, CA 94720-7360. Members are listed at the end of the manuscript.

## Abstract

**Background:** Long COVID, also known as post-acute sequelae of COVID-19 (PASC), is a poorly understood condition with symptoms across a range of biological domains that often have debilitating consequences. Some have recently suggested that lingering SARS-CoV-2 virus in the gut may impede serotonin production and that low serotonin may drive many Long COVID symptoms across a range of biological systems. Therefore, selective serotonin reuptake inhibitors (SSRIs), which increase synaptic serotonin availability, may prevent or treat Long COVID. SSRIs are commonly prescribed for depression, therefore restricting a study sample to only include patients with depression can reduce the concern of confounding by indication.

**Methods:** In an observational sample of electronic health records from patients in the National COVID Cohort Collaborative (N3C) with a COVID-19 diagnosis between September 1, 2021, and December 1, 2022, and pre-existing major depressive disorder, the leading indication for SSRI use, we evaluated the relationship between SSRI use at the time of COVID-19 infection and subsequent 12-month risk of Long COVID (defined by ICD-10 code U09.9). We defined SSRI use as a prescription for SSRI medication beginning at least 30 days before COVID-19 infection and not ending before COVID-19 infection. To minimize bias, we estimated the causal associations of interest using a nonparametric approach, targeted maximum likelihood estimation, to aggressively adjust for high-dimensional covariates.

**Results:** We analyzed a sample (*n* = 506,903) of patients with a diagnosis of major depressive disorder before COVID-19 diagnosis, where 124,928 (25%) were using an SSRI. We found that SSRI users had a significantly lower risk of Long COVID compared to nonusers (adjusted causal relative risk 0.90, 95% CI (0.86, 0.94)).

**Conclusion:** These findings suggest that SSRI use during COVID-19 infection may be protective against Long COVID, supporting the hypothesis that serotonin may be a key mechanistic biomarker of Long COVID.

## BACKGROUND

COVID-19 infection can have debilitating long-term consequences. Long COVID, also known as post-acute sequelae of COVID-19 (PASC), includes symptoms across a range of biological systems that can occur following COVID-19 infection. Millions of adults in the United States may be experiencing Long COVID, the majority of whom only experienced mild to moderate COVID-19 infection.^1,2^ Even though more than 10% of COVID-19 patients develop PASC, we have few insights regarding options for treatment and prevention.^3^ Insights regarding treatments that may prevent Long COVID are crucial to preventing this condition and understanding its etiology.

Investigators have hypothesized several biological mechanisms that drive Long COVID and lead to clusters of symptoms. These explanations include (1) persistent COVID-19 viral load, (2) chronic hyperinflammation, (3) platelet and coagulation issues, and (4) central nervous system dysfunction.^4,5^ Previous studies have clustered these symptoms and speculated that these pathways may be distinct disorders caused by different components of acute COVID-19 infection.^6^ On the other hand, recent investigations have highlighted reduced serotonin as a driver of all four of these symptom clusters.^4^ A metabolomics investigation found that persistent COVID-19 viral load led to sustained interferon response, decreased uptake of tryptophan (a serotonin precursor), hypercoagulation, and subsequent decrease in serotonin.^4^ This peripheral serotonin deficiency leads to reduced vagus nerve activity, which subsequently contributes to decreased hippocampal activity, which can result in memory loss and cognitive dysfunction.^4^

SSRIs are the first-line medication class used to treat depression. They have high tolerability and are considered safe and effective.^7,8^ SSRI’s mechanism of action is to prevent serotonin reuptake by inhibiting serotonin transporter at the presynaptic axon terminal. The prevention of this reuptake allows for a greater concentration of serotonin in the synaptic cleft that can bind to receptors.^7^ Compared with other classes of antidepressants, such as tricyclic antidepressants or monoamine oxidase inhibitors, SSRIs have fewer side effects due to fewer effects on other neurotransmitters and receptors.^7^ Given SSRI’s specific targeting of serotonin, it is an ideal candidate to evaluate the role of serotonin in the development of Long COVID.

Several studies have investigated the relationship between SSRI use and acute COVID-19 infection as well as Long COVID. The TOGETHER trial found that early treatment with the SSRI fluvoxamine improved COVID-19 patient recovery.^9^ On the other hand, the COVID-OUT trial found that fluvoxamine treatment during acute COVID-19 infection did not reduce the cumulative incidence of Long COVID (1.36, 95% CI (0.78–2.34)), although this analysis included a relatively small sample size (334 patients assigned to fluvoxamine) and may have been underpowered.^10^ More broadly, previous studies have found that SSRI use may reduce the probability of hospitalization or mortality due to COVID-19 infection.^11,12^ A 2022 study evaluated the relationship between SSRI use and the predicted PASC and found that SSRI use was associated with 0.75 (95% CI, 0.62, 0.90) times the risk of predicted PASC compared to non-use.^13^ While this observational study provided evidence that SSRI use may be protective against Long COVID, this study used predicted PASC diagnosis (via XGboost machine learning) as its primary endpoint, rather than an actual PASC diagnosis. This predicted PASC status did not directly include SSRI use in its prediction, but it did use a myriad of other diagnoses and medications, which may be correlated with SSRI use and may have induced bias. Furthermore, the study used a general sample of SSRI users and non-users rather than restricting to conditions that may yield SSRI use, leading to the possibility of residual confounding by indication. In addition, a recent observational study found that patients experiencing Long COVID experienced improvement in self-reported symptoms following treatment with SSRIs.^14^

Several studies to date have evaluated the impact of individual types of SSRIs on Long COVID. A multi-system study of the relationship between serotonin and Long COVID hypothesized that fluoxetine may be particularly effective in preventing and treating Long COVID, and they found that treating mice with fluoxetine improved cognitive function and restored tryptophan levels.^4^ Furthermore, animal models involving fruit flies have demonstrated that the specific SSRI types fluoxetine, escitalopram, citalopram, and paroxetine may differentially impact serotonin reuptake.^15^ A systematic review of studies evaluating the use of fluvoxamine for COVID-19 and Long COVID suggested that baseline use of fluvoxamine may reduce the risk of Long COVID due to the drug’s sigma 1 receptor agonist activity and the role of sigma 1 receptor activity in acute COVID-19 infection.^16^ Observational analyses of human electronic health record (EHR) data did not find a significant difference in the relationship between moderate to high-affinity sigma 1 receptor agonist SSRIs (fluvoxamine, fluoxetine, escitalopram, and citalopram) versus non-high affinity SSRIs (sertraline and paroxetine) in their impact on Long COVID.^13^

Identifying interventions that prevent Long COVID is crucial for clinical applications as well as our understanding of underlying biological mechanisms that cause Long COVID. Nationally sampled EHR databases, such as the National COVID Cohort Collaborative (N3C), provide an excellent opportunity to evaluate these hypotheses but require analytic methods that can aggressively adjust for high-dimensional confounders without making bias-inducing parametric assumptions.^17–22^ While randomized controlled trials may eventually provide definitive evidence regarding the benefit of SSRI use to prevent or treat Long COVID, observational analyses, using appropriate methods that are designed to leverage the complexity, including missing data, and large sample sizes characteristic of EHR real-world data (RWD) can provide early insights regarding the relationship between SSRI use and Long COVID. Thus, to evaluate the relationship between SSRI use during COVID-19 infection and Long COVID cumulative incidence, we conducted an observational analysis of individuals in N3C with an acute COVID-19 infection and comorbid depression diagnosis using a machine-learning-based method targeted to reduce bias due to confounding and missing data (Targeted Machine Learning).^17,18,20–22^

**Figure 1.**
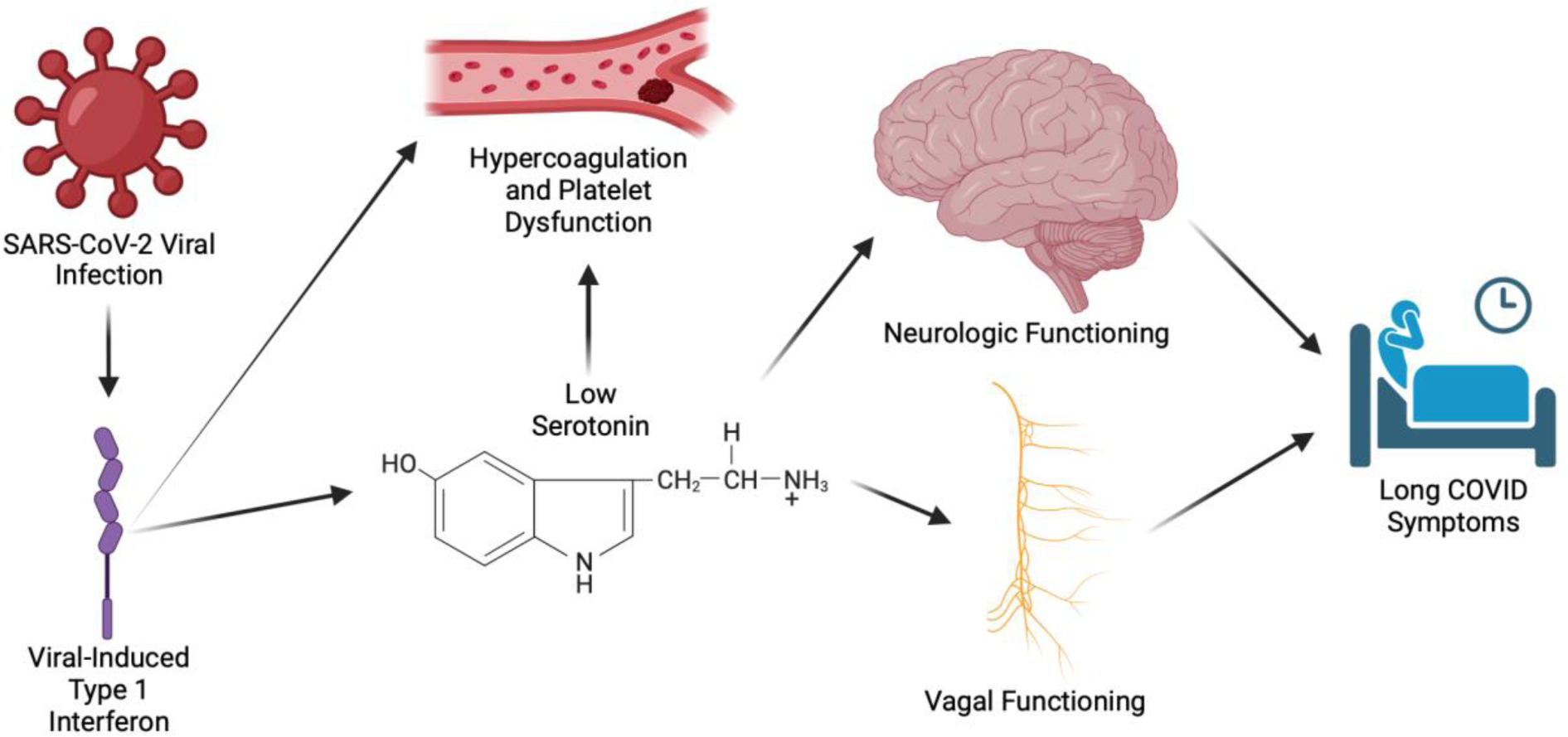
Hypothesized mechanism of the relationship between serotonin and Long COVID^4,23^

## METHODS

### Study sample, data source, and study design

Our primary study sample included individuals with a diagnosis of acute COVID-19 infection between September 1, 2021 (PASC ICD-10 code U09.9 released October 1, 2021) and December 1, 2022, as well as a comorbid diagnosis of major depressive disorder.^24^ This sample was drawn from patients in N3C (DUR-80D09B6), which includes 22 million patients from 82 healthcare institutions.^19^ N3C provides high-dimensional data on these participants, which enables researchers to conduct evaluations of a wide range of factors related to Long COVID and acute COVID-19 infection while rigorously adjusting for factors related to medical history and sociodemographic information.

We constructed a retrospective cohort of patients in N3C who were diagnosed with major depressive disorder (depression) before their acute COVID-19 infection. As SSRI prescription is often indicated by depression, we restricted our sample to only include those with depression to limit confounding by indication. We evaluated SSRI use (as a time-invariant, binary variable) at the time of acute COVID-19 infection, and we assessed patients’ cumulative incidence of Long COVID (PASC) between 1 and 12 months following acute COVID-19 infection, comparing SSRI users to nonusers. We included patients from 80 data partners (contributors of patient data) in N3C. We found that 23% of data partners did not report PASC diagnosis, and 6% of data partners did not report SSRI use in this study sample.

N3C identified patients with acute COVID-19 infection as patients who had either (1) at least one laboratory test with a positive result, (2) at least one “strong positive” diagnostic code in ICD-10 or SNOMED, (3) at least two “weak positive” diagnostic codes in ICD-10 or SNOMED.^25^ N3C defines the acute COVID-19 infection date as the date of the first positive diagnostic code or laboratory rest.

### Key covariates

#### Exposures

We defined the exposure of interest as a binary variable that represents SSRI use (fluoxetine, sertraline, paroxetine, citalopram, escitalopram, fluvoxamine, and vilazodone, phenotyped using RxNorm) during incident COVID-19 infection. We defined SSRI users as patients who began using an SSRI at least 30 days before COVID-19 infection and continued through acute COVID-19 infection (binary, time-invariant), and we defined all other patients as nonusers.

#### Outcomes

Our outcome of interest was observed PASC diagnosis, which was defined by ICD code U09.9, between 1 and 12 months following acute COVID-19 infection.^26^ We included observed PASC (U09.9) diagnosis as our outcome of interest, as it provides a standardized metric of Long COVID incidence across diagnostic settings. In contrast, using the predicted probability of PASC diagnosis (e.g. via machine learning methods) may induce bias if the predictions are generated using the same EHR data as the exposures of interest.^13^ We ensured that all participants would have 12 months of follow-up by restricting to participants who were diagnosed with COVID-19 infection between September 1, 2021 (1 month before the implementation of ICD code U09.9),^24^ and December 1, 2022, and including PASC diagnosis data within 12 months of COVID-19 infection (i.e. up to December 1, 2023). We will describe PASC (ICD code U09.9) as “Long COVID” hereafter.

#### Subgroups of interest

We created subgroups of individuals with specific SSRI drug type exposures for SSRIs with a sufficient sample size, which included fluoxetine, sertraline, paroxetine, citalopram, and escitalopram. Vilazodone and fluvoxamine had insufficient sample size and therefore were excluded from subgroup analyses. We constructed separate models for each SSRI of interest to assess potential effect heterogeneity. Furthermore, we conducted exploratory analyses of potential dose-response relationships by analyzing subgroups defined by SSRI dosage among fluoxetine users, given fluoxetine’s large sample size and hypothesized relationship with Long COVID.^13^

#### Confounders and other covariates

We were able to extract extensive medical histories from patients in N3C to adjust for a rich history of patient data and thus avoid unmeasured confounding. Our set of baseline covariates included the following: healthcare utilization rate (number of visits pre-COVID-19 infection and healthcare visits per month before COVID-19 infection), sex, age at acute COVID-19 infection, race/ ethnicity, common data model format, region of residence, body mass index (BMI), tobacco smoking status, obesity, diabetes, chronic lung disease, heart failure, hypertension, use of systemic corticosteroids, depression severity, bipolar disorder, whether the patient was immunocompromised, and the number of COVID-19 vaccination doses before infection.^27^ We defined a healthcare visit as a single visit, or cluster of visits, to a healthcare provider that was associated with a given medical condition, diagnosis, or procedure. We included county-level socioeconomic variables that included the percent of the county with an income level below the poverty line and the county’s social deprivation index score. We also used methods that can minimize bias due to differential monitoring among patients, by including an indicator variable for whether the patient had a documented healthcare visit between 1 and 12 months following acute COVID-19 infection (the outcome observation period). We have included additional details regarding covariates in Appendix 1.

#### Negative control outcome

We sought to evaluate a negative control outcome to evaluate the possibility of bias in the underlying data. We evaluated bone fracture diagnosis between 1 and 12 months after acute COVID-19 diagnosis as a negative control outcome.

### Analysis

To accomplish the goals of using nonparametric statistical methods that could adjust for rich, messy patient history and monitoring data, we applied a Targeted Learning approach, which is well-suited for this context of observational analyses of electronic health record data.^17,18,21,28^ Traditional parametric analyses make assumptions regarding model form and relationships between covariates, and these assumptions will inevitably be violated in this high-dimensional setting. This potential model misspecification would increase bias and the probability of Type 1 error, particularly given our large sample size.^17,18,21,28^ On the other hand, Targeted Learning (TL) utilizes advances in machine learning and causal inference by capitalizing on the extensive data to minimize bias introduced by arbitrary modeling assumptions, which can result in improper under-adjustment of confounders. In addition, TL methods provide robust statistical inference despite data-adaptive, machine-learning methods being used to estimate the statistical relationships of interest.

Our goal was to estimate the impact of SSRI use at the time of COVID-19 infection on the probability of developing Long Covid by comparing the predicted distribution of Long Covid under universal versus no use of SSRIs among our target population, patients with major depressive disorder, under a scenario of universal monitoring of patients between 1 and 12 months after acute COVID-19 infection. Our analysis approach first used Super Learner, an ensemble machine learning algorithm, to predict Long COVID status given individual covariate status (diagnoses, treatment, demographics, and other history).^29,30^ Super Learner uses cross-validation to determine the optimal weighting of candidate algorithms to maximize a parameter of interest. Next, we used Targeted Maximum Likelihood Estimation to estimate the causal parameter of interest (the risk ratio) comparing Long COVID incidence in the exposed versus unexposed population.^17,18,21,28^ Targeted Maximum Likelihood Estimation allows us to generate interpretable measures of association, such as a risk ratio while reducing bias. In addition, Targeted Maximum Likelihood Estimation is doubly robust, meaning that it guarantees consistent estimation as long as the outcome regression or propensity score is estimated consistently.^17,18,21,28^

As Super Learner guarantees that the ensemble will perform at least as well as the best-performing candidate learner, given sufficient sample size, we sought to include a diverse library of parametric and nonparametric candidate algorithms to ensure optimal performance.^29,30^ We included the following candidate algorithms: generalized linear models (SL.glm), Bayesian Additive Regression Trees (tmle.SL.dbarts2), Generalized Linear models net (SL.glmnet), XGBoost (SL.xgboost), Caret (SL.caret), Caret Recursive Partitioning and Regression Trees (SL.caret.rpart), K Nearest Neighbors (SL.knn), Neural Net (SL.nnet), Random Forest (SL.randomForest), and Recursive Partitioning and Regression Trees (SL.rpart).^29,30^ We also used cross-validated (cross-fitted) Targeted Maximum Likelihood Estimation (TMLE), which avoids overfitting and adds robustness.^17,18,21,28^

We applied a *W*, *A*, *Δ*, *ΔY* data structure, where *W* referred to our baseline confounders and covariates of interest, *A* referred to our exposure of interest, *Δ* referred to participant observation during the outcome period (months 1-12), and *ΔY* referred to our observed outcome. If a participant did not have a healthcare visit during the outcome window (months 1-12 following COVID-19 infection), which could be due to lack of healthcare engagement or patient death before observation, Delta would be equal to 0. We defined our causal parameter of interest as *E*(*Y*(1,1) − *Y*(0,1)), where *Y*(*a*, *Δ* = 1) is defined as the counterfactual outcome if SSRI status is set to *A=a*, and the person was monitored during the at-risk period (*Δ* = 1). We intervened on *Δ* to ensure that all participants were observed (had at least one healthcare visit) during the outcome window (between 1 and 12 months following COVID-19 infection). We make the assumption that the subset of confounders that are observed for each subject was sufficient to adjust for confounding; operationally, this was done by adding new basis functions for confounders with missing values, which were indicators that the variable was observed, and imputed values for the missing covariate. This allows us to aggressively adjust for confounding and keep observations with missing covariate information (*W*).^22,27^

#### Sensitivity analyses

In order to evaluate underlying biases in our analysis and data, we conducted a nonparametric sensitivity analysis.^31^ This nonparametric sensitivity analysis allows us to compare the theoretical bias that would nullify our results to benchmarks, such as the difference between our observed adjusted estimate and unadjusted estimate, that could explain the magnitude of our observed association. Furthermore, we evaluated the relationship between SSRI use during COVID-19 infection and bone fracture between 1 and 12 months following COVID-19 infection as a negative control outcome analysis. We compared our observed, adjusted result to the (1) unadjusted association and (2) the negative control outcome association.

## RESULTS

### Descriptive statistics

We analyzed electronic health records from a sample of 506,903 patients who were diagnosed with major depressive disorder before COVID-19 diagnosis. Among these patients, 124,928 (25%) were using an SSRI at the time of COVID-19 infection and 381,975 (75%) were not (Table 1). We found that SSRI users generally had a greater burden of disease and more markers of poor health than SSRI non-users. Among SSRI users, 17% were morbidly obese compared to 15% of non-users,13% had experienced heart failure compared to 10% of non-users, 35% had experienced lung disease compared to 30% of nonusers, 6% were diagnosed with bipolar disorder compared to 5% of nonusers, 19% were smokers compared to 17% of nonusers, and 64% used systemic corticosteroids compared to 49% of non-users. We also found that 9% of SSRI users had severe major depressive disorder, compared with 7% of non-SSRI users. SSRI users had a healthcare utilization rate of 2.9 healthcare visits per month, while nonusers had 2.5 healthcare visits per month. We found that 30% of SSRI users had at least one dose of a COVID-19 vaccination, and 34% of nonusers had at least one vaccination dose.

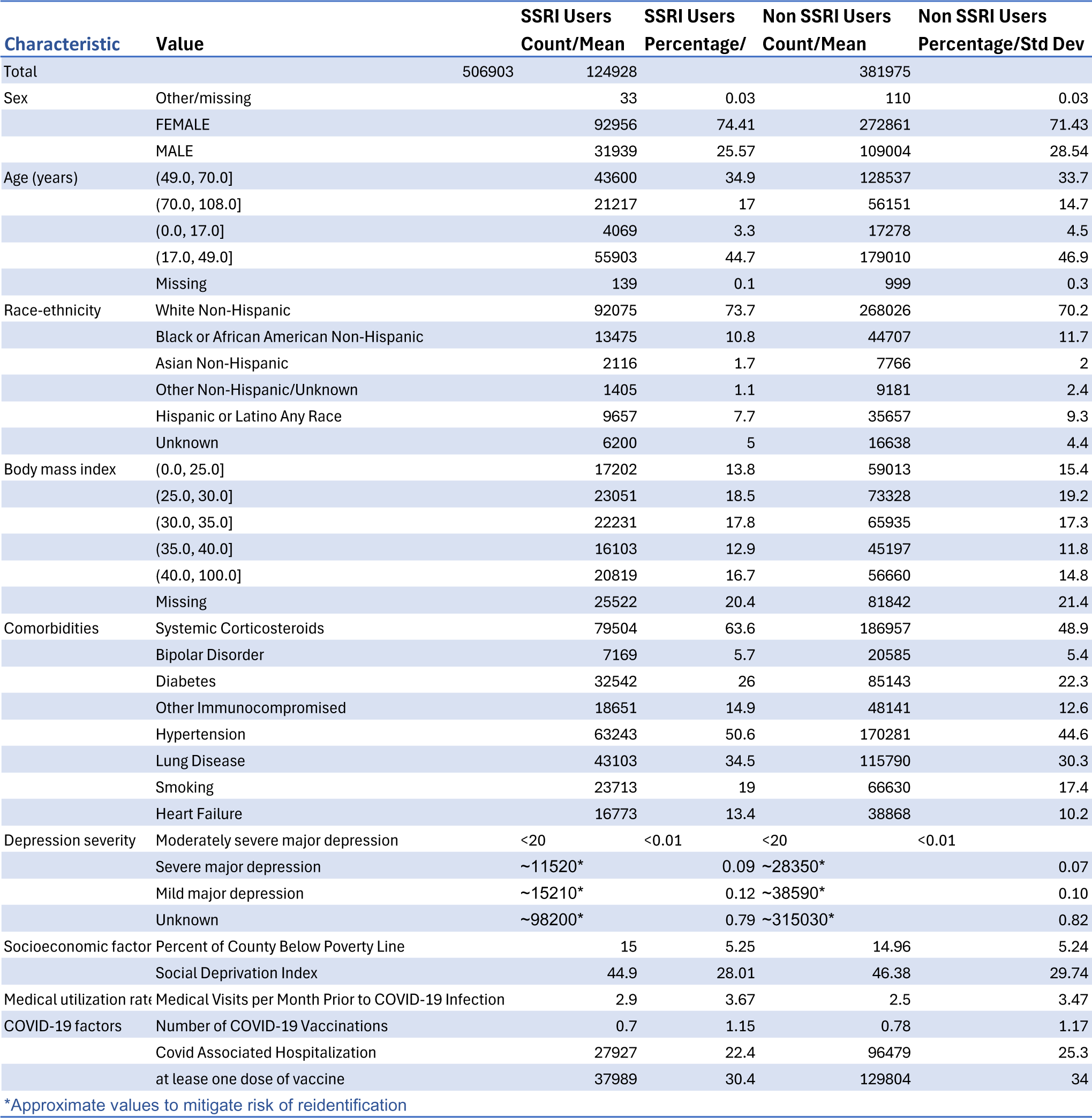

### Relationship between SSRI use and Long COVID

We found that SSRI users had a lower risk of Long COVID (adjusted risk ratio (aRR) 0.901, 95% confidence interval (CI) (0.861, 0.943)) compared to nonusers (Figure 2). Adjustment for baseline confounders shifted the estimate a fair distance from the unadjusted association, which indicated a positive relationship between SSRI use and Long COVID (unadjusted RR 1.231, 95% CI (1.183, 1.280)) (Table 2).

**Figure 2.**
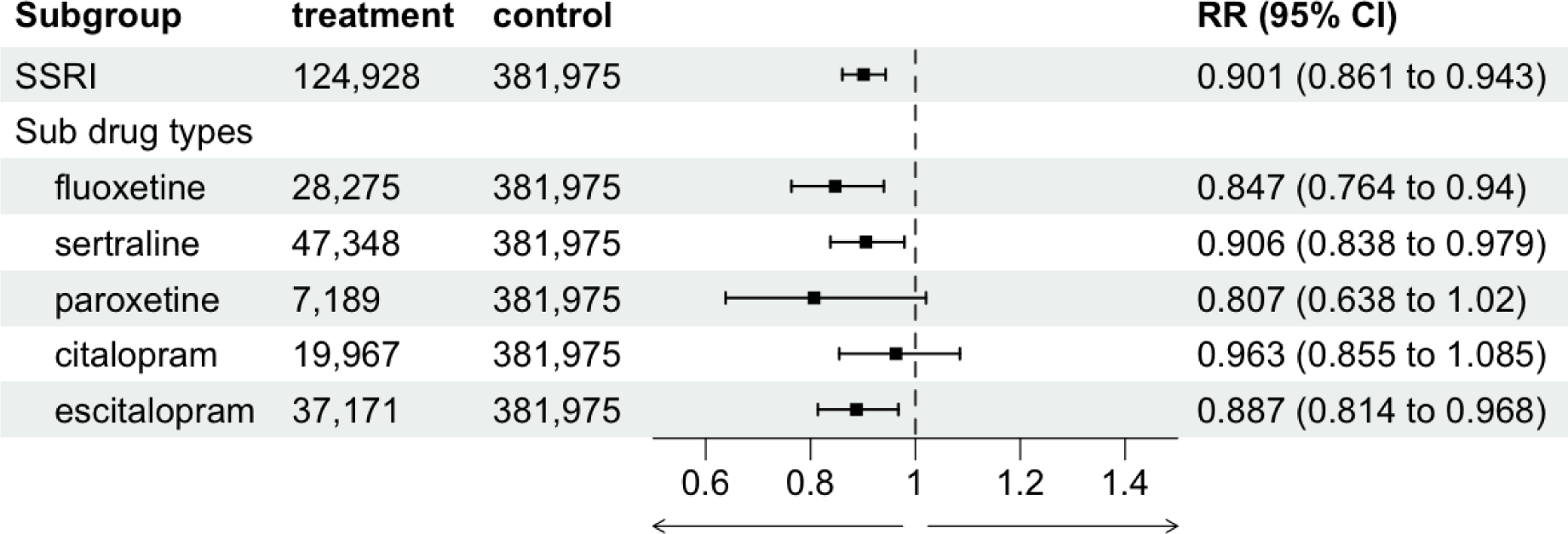
Relationship between SSRI use (overall and by SSRI type) and Long COVID among patients with depression.

**Table 2.** Relationships between SSRI use during acute COVID-19 infection and 12-month Long COVID risk.

| SSRI Type | Sample size Using Drug | Sample size not using drug | Unadjusted risk using drug | Unadjusted Risk not using Drug | Unadjusted RR | CI (lower) | CI (upper) | Adjusted risk using drug | Adjusted Risk not using drug | Adjusted RR | (95% CI) |
| --- | --- | --- | --- | --- | --- | --- | --- | --- | --- | --- | --- |
| Any SSRI | 124928 | 381975 | 0.027 | 0.0217 | 1.2306 | 1.1827 | 1.2804 | 0.0211 | 0.0234 | 0.9010 | (0.8607, 0.9433) |
| Fluoxetine | 28275 | 381975 | 0.027 | 0.0217 | 1.2557 | 1.1675 | 1.3506 | 0.0187 | 0.0221 | 0.8471 | (0.7636, 0.9398) |
| Sertraline | 47348 | 381975 | 0.026 | 0.0217 | 1.1891 | 1.1206 | 1.2617 | 0.0203 | 0.0224 | 0.9058 | (0.8378, 0.9792) |
| Paroxetine | 7189 | 381975 | 0.027 | 0.0217 | 1.2315 | 1.0694 | 1.4182 | 0.0176 | 0.0219 | 0.8068 | (0.6379, 1.0204) |
| Citalopram | 19967 | 381975 | 0.028 | 0.0217 | 1.3048 | 1.1996 | 1.4191 | 0.0213 | 0.0221 | 0.9630 | (0.8548, 1.0849) |
| Escitalopram | 37171 | 381975 | 0.027 | 0.0217 | 1.2653 | 1.1867 | 1.3491 | 0.0198 | 0.0223 | 0.8875 | (0.8140, 0.9676) |

We evaluated the relationship between individual SSRI types and Long COVID (fluoxetine, sertraline, paroxetine, escitalopram, and citalopram). In our subgroup analysis, comparing the use of each of the five SSRIs to no SSRI use, we found that fluoxetine (aRR 0.847, 95% CI (0.763, 0.940)), sertraline (aRR 0.905, 95% CI (0.0.838, 0.979), and escitalopram (aRR 0.887, 95% CI (0.814, 0.967)) were associated with a lower cumulative incidence of Long COVID. We did not detect an association between the use of paroxetine (aRR 0.807, 95% CI (0.638, 1.020)) or citalopram (aRR 0.963, 95% CI (0.855, 1.085)) and the risk of Long COVID. We did not find evidence of a dose-response relationship between fluoxetine dose and risk of Long COVID (60 mg vs. 10 mg aRR 1.421, 95% CI (0.656, 3.080)) (Supplemental Table 1).

### Sensitivity analyses and confounding

We found that the relationship between SSRI use and Long COVID was strongly and qualitatively confounded, as the unadjusted estimate indicated a positive (harmful) correlation, but the adjusted estimate indicated a negative (protective) correlation. We observed the change in estimate following the backward exclusion of each covariate, where we defined “confounder RR” as the risk ratio in the fully-adjusted model divided by the risk ratio of the partially-adjusted model (with the covariate excluded) (Supplemental Table 2). We found that the strongest confounders of the relationship between SSRI use and Long COVID were baseline systemic corticosteroid use (confounder RR 0.983), monitoring during the outcome window (binary indicator of healthcare visitation between 1 and 12 months after acute COVID-19) (confounder RR 0.989), number of healthcare visits at baseline (confounder RR 0.995), and social deprivation index (confounder RR 1.005). We also evaluated the impact of excluding two groups of covariates, healthcare utilization (number of healthcare visits before baseline, healthcare visitation rate before baseline, and monitoring during the outcome window) and baseline general health and comorbidities general health and comorbidities (BMI, chronic lung disease, diabetes, obesity, immunocompromised status, smoking, corticosteroid use, hypertension, and COVID-19 vaccinations). We found that excluding factors related to healthcare utilization led to a confounder RR of 0.969 while excluding factors related to baseline comorbidities and health led to a confounder RR of 0.979.

We conducted a nonparametric sensitivity analysis to evaluate the potential impact of bias on our results (Figure 3). We found that 0.33 units of bias, where one unit corresponds to the difference between our adjusted and unadjusted estimate, could lead to a value as extreme as our observed estimate, due to random variation alone.

**Figure 3.**
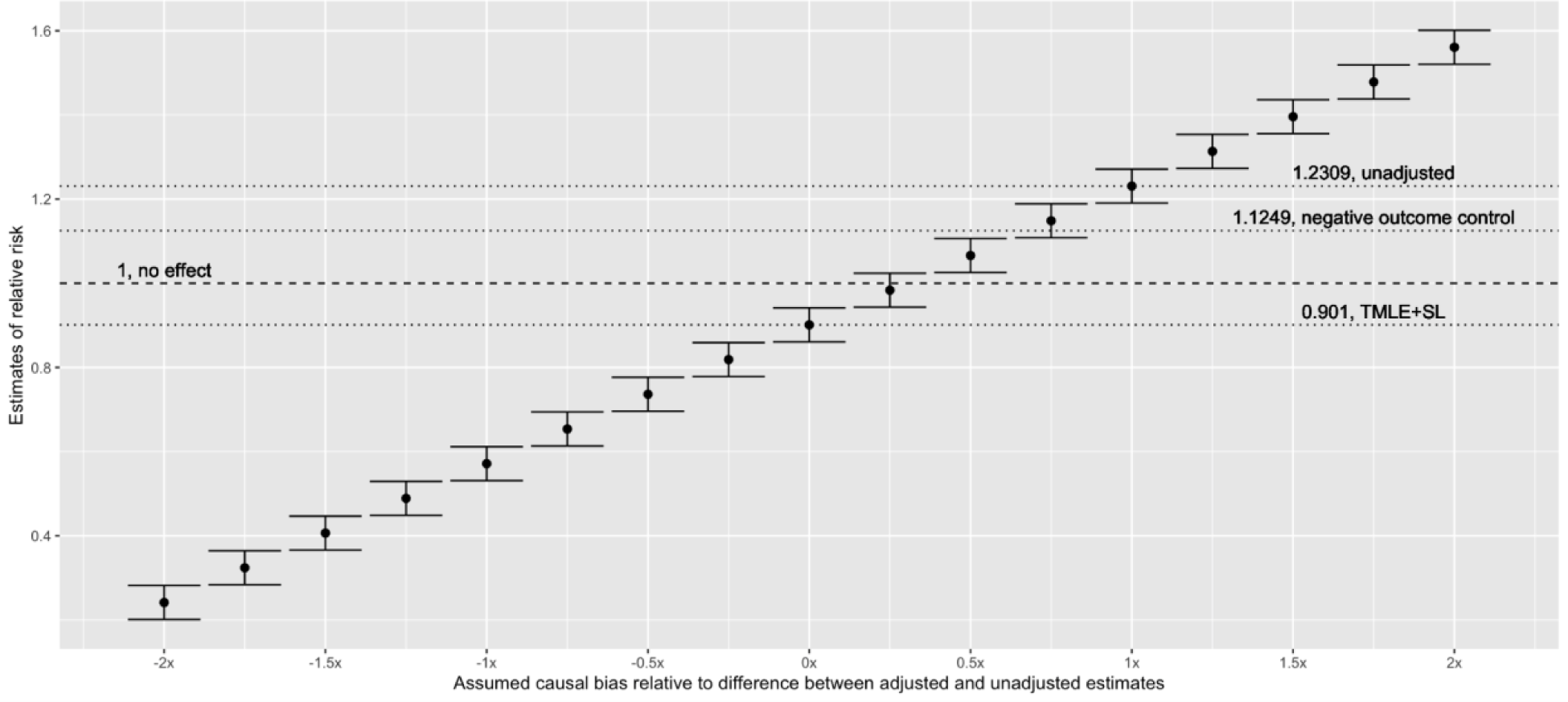
Nonparametric sensitivity analysis depicting the observed, adjusted risk ratio (TMLE+SL) as well as the unadjusted risk ratio (unadjusted) and the results of an analysis of a negative control outcome (bone fracture).

## DISCUSSION

We found a protective effect of SSRI use at the time of acute COVID-19 infection on subsequent 12-month risk of Long COVID. These results are consistent with the hypothesis that SSRIs may be an effective intervention to prevent Long COVID, which also supports the hypothesis that serotonin may play a role in the development of Long COVID. Randomized controlled trials are currently underway to evaluate the ability of SSRIs to prevent or treat Long COVID (NCT05874037, NCT06128967). With ongoing COVID-19 transmission, the risk of Long COVID remains prevalent, and finding interventions to prevent Long COVID remains prudent. SSRIs may serve as an important tool in preventing this condition and limiting the rippling effects of the COVID-19 pandemic.

Our findings regarding the protective effect of SSRI use on Long COVID risk are consistent with previous studies. This observed treatment effect, a risk ratio of 0.90 (95% CI 0.86, 0.94), is less protective than a previous analysis, which found a risk ratio of 0.76 (95% CI 0.62, 0.90).^13^ The difference in these effects may be attributed to several potential factors, including the previous study’s use of predicted Long COVID status rather than observed diagnosis (yielding a prevalence of 15% rather than 2%) as well as our restriction to only include patients with a diagnosis of major depressive disorder.^13^ These considerations may avoid bias and confounding due to indication, respectively.

The observed unadjusted and adjusted estimates varied qualitatively (from harmful to protective). The unadjusted association indicating a harmful relationship between SSRI use and Long COVID may be explained by strong confounding due to various factors and is supported by our finding of imbalance and confounding due to healthcare utilization, medication usage, and socioeconomic factors. This phenomenon is referred to as Simpson’s paradox, where findings can vary drastically when causal inference considerations are taken into account.^32^ Our nonparametric sensitivity analysis contextualizes these relationships and indicates that, for our observed, adjusted estimate to be a Type 1 error, we would need to have 0.33 units of bias, where one unit refers to the difference between adjusted and unadjusted estimates.^31^ The qualitative shift in our observed measure of association following multivariate adjustment underscores the need for additional research in this area. Furthermore, our finding that factors related to healthcare utilization rate (number of healthcare visits before baseline and outcome monitoring indicator) were strong confounders of our observed relationship highlights the importance of addressing differential healthcare utilization rates and other causal considerations in observational studies that rely on Long COVID diagnosis as an outcome of interest.^27^

These findings provide additional support for the hypothesis that low serotonin is a driver of Long COVID incidence and that SSRIs may prevent or treat Long COVID. Although SSRI treatment may not directly intervene on remnants of the SARS-CoV-2 virus, which leads to the sustained release of viral RNA-induced type 1 interferons, which decreases tryptophan uptake and prevents cortisol production, SSRI use may still interrupt this causal pathway of disease etiology.^4^ This potential biological pathway may include the increase of synaptically available serotonin, through inhibited serotonin reuptake, which increases vagal signaling to prevent neurological and cognitive symptoms of Long COVID.^7^ As this hypothesis posits that low serotonin is a downstream effect of lingering SARS-CoV-2 virus and sustained interferon 1 response, these findings also hint at interventions that aim to detect or treat persistent viral load of SARS-CoV-2 or viral RNA-induced type 1 interferon.

We did not find evidence of heterogeneity of the relationship between SSRI use and Long COVID depending on SSRI type. This finding is consistent with a previous study, which did not find a differential impact of moderate to high-affinity sigma 1 receptor agonist SSRIs (fluvoxamine, fluoxetine, escitalopram, and citalopram) versus non-high affinity SSRIs (sertraline and paroxetine) in their relationship with Long COVID.^13^ We caution readers to consider this finding in the context of a few limitations. Residual confounding due to indication, as different depressive symptomatology, comorbidities, side effects, and tolerance may lead providers to prescribe a specific SSRI over another SSRI. For instance, citalopram and paroxetine are often prescribed for obsessive-compulsive disorder, which may be associated with Long COVID symptoms.^33–36^ Furthermore, our analysis of paroxetine was limited by a small sample size of users (*n* = 7,189). We also did not find evidence of a dose-response relationship between fluoxetine and Long COVID. This may be explained due to a large proportion of missingness of dose information leading to a small functional sample size. Future studies should further explore the possibility of a dose-response relationship.^8^

### Strengths and Limitations

A strength of this study is its large, national sample size of participants and the broad range of high-dimensional data that we included via N3C. This rich data source allows us to construct a cohort of patients with a diagnosis of major depressive disorder, assess their SSRI use at the time of COVID-19 infection, and evaluate their probability of Long COVID diagnosis. Furthermore, the documentation of comorbidities, sociodemographic information, and other medical history allows for rigorous multivariate adjustment.

A second strength of this study is the analytic methods that we applied. A Targeted Learning approach, involving Super Learner and targeted maximum likelihood estimation, allows for efficient and flexible estimation while making minimal parametric assumptions.^17,18,21,29,37^ With this large sample size of high-dimensional data, this allows us to aggressively reduce bias due to measured, potentially high-dimensional confounding and to do so with nearly no model assumptions. Furthermore, these methods allowed us to intervene on participant observation during the outcome window, which is an important driver of differential outcome ascertainment.^27,31^ Finally, nonparametric sensitivity analyses allowed us to determine the extent to which our results are vulnerable to bias. Cumulatively, these methods provide a replicable framework for investigators to conduct rigorous observational analyses using electronic health record databases such as N3C.

A third strength of this study was its ability to flexibly account for and intervene on the missingness of the outcome and heterogeneous monitoring.^31^ There is significant heterogeneity in N3C’s documentation of Long COVID diagnoses (our outcome of interest), as is common with electronic health record databases. Previous studies have found that Long COVID diagnosis is strongly correlated with healthcare utilization rate.^27,38^ We sought to control for healthcare utilization rate by adjusting for multiple factors related to healthcare utilization, including healthcare visits per month before COVID-19 infection. In addition, we were able to use novel causal inference framing to define our parameter of interest at the ratio of probabilities of PASC under universal monitoring, i.e., by “intervening” on whether an individual had a healthcare visit between 1 and 12 months following acute COVID-19 infection (the period at-risk for Long COVID), to observe the counterfactual impact of SSRI exposure given that all participants were observed during the period at-risk for the outcome.^39,40^ It remains possible that residual confounding due to healthcare utilization rate remains, although this would likely bias our estimate towards the null, indicating that our observed measure of association is likely conservative.^27^

This study had several limitations. We defined the exposure of interest as a binary, time-invariant variable based on SSRI use during COVID-19 infection. It remains possible that factors related to the duration of SSRI use, timing of SSRI use, or SSRI dosage may modify this relationship and should be explored in a future study. Furthermore, PASC diagnosis (ICD code U09.9) has limited sensitivity and may fail to reflect heterogeneity within Long COVID subtypes (e.g. neurological versus gastrointestinal symptoms). Finally, as an observational study, this analysis may be subject to residual bias and investigators should conduct randomized controlled trials to corroborate these findings. The generalizability of N3C patients has been described previously.^6,38,41^ N3C is a broad, national sample of participants, as it relies on electronic health record data, but it skews towards participants who engage more with healthcare systems. This yields a study population that is generally older, has more comorbidities than the general population, and underrepresents un- or underinsured patients.^42^

## CONCLUSION

This study suggests that the use of SSRIs during acute COVID-19 infection is associated with a lower risk of Long COVID among patients with major depressive disorder. These results support the hypothesis that serotonin is a key mechanistic biomarker of Long COVID and that SSRIs may be an effective intervention to prevent Long COVID.

## Data Availability

All data in the present study are available upon request from the National COVID Cohort Collaborative (N3C).

https://covid.cd2h.org

## N3C Attribution

The analyses described in this manuscript were conducted with data or tools accessed through the NCATS N3C Data Enclave https://covid.cd2h.org and N3C Attribution & Publication Policy v 1.2-2020-08-25b supported by NCATS U24 TR002306, Axle Informatics Subcontract: NCATS-P00438-B, and the Bill & Melinda Gates Foundation: OPP1165144. This research was possible because of the patients whose information is included within the data and the organizations (https://ncats.nih.gov/n3c/resources/data-contribution/data-transfer-agreement-signatories) and scientists who have contributed to the on-going development of this community resource [https://doi.org/10.1093/jamia/ocaa196].

## Disclaimer

The N3C Publication committee confirmed that this manuscript (MSID:1784.118) is in accordance with N3C data use and attribution policies; however, this content is solely the responsibility of the authors and does not necessarily represent the official views of the National Institutes of Health or the N3C program.

## IRB

The N3C data transfer to NCATS is performed under a Johns Hopkins University Reliance Protocol # IRB00249128 or individual site agreements with NIH. The N3C Data Enclave is managed under the authority of the NIH; information can be found at https://ncats.nih.gov/n3c/resources.

This research project was approved by the University of California, Berkeley Committee for the Protection of Human Subjects (CPHS protocol number 2022-01-14980). This approval is issued under University of California, Berkeley Federalwide Assurance #00006252.

## Individual Acknowledgements For Core Contributors

We gratefully acknowledge the following core contributors to N3C: Adam B. Wilcox, Adam M. Lee, Alexis Graves, Alfred (Jerrod) Anzalone, Amin Manna, Amit Saha, Amy Olex, Andrea Zhou, Andrew E. Williams, Andrew Southerland, Andrew T. Girvin, Anita Walden, Anjali A. Sharathkumar, Benjamin Amor, Benjamin Bates, Brian Hendricks, Brijesh Patel, Caleb Alexander, Carolyn Bramante, Cavin Ward-Caviness, Charisse Madlock-Brown, Christine Suver, Christopher Chute, Christopher Dillon, Chunlei Wu, Clare Schmitt, Cliff Takemoto, Dan Housman, Davera Gabriel, David A. Eichmann, Diego Mazzotti, Don Brown, Eilis Boudreau, Elaine Hill, Elizabeth Zampino, Emily Carlson Marti, Emily R. Pfaff, Evan French, Farrukh M Koraishy, Federico Mariona, Fred Prior, George Sokos, Greg Martin, Harold Lehmann, Heidi Spratt, Hemalkumar Mehta, Hongfang Liu, Hythem Sidky, J.W. Awori Hayanga, Jami Pincavitch, Jaylyn Clark, Jeremy Richard Harper, Jessica Islam, Jin Ge, Joel Gagnier, Joel H. Saltz, Joel Saltz, Johanna Loomba, John Buse, Jomol Mathew, Joni L. Rutter, Julie A. McMurry, Justin Guinney, Justin Starren, Karen Crowley, Katie Rebecca Bradwell, Kellie M. Walters, Ken Wilkins, Kenneth R. Gersing, Kenrick Dwain Cato, Kimberly Murray, Kristin Kostka, Lavance Northington, Lee Allan Pyles, Leonie Misquitta, Lesley Cottrell, Lili Portilla, Mariam Deacy, Mark M. Bissell, Marshall Clark, Mary Emmett, Mary Morrison Saltz, Matvey B. Palchuk, Melissa A. Haendel, Meredith Adams, Meredith Temple-O’Connor, Michael G. Kurilla, Michele Morris, Nabeel Qureshi, Nasia Safdar, Nicole Garbarini, Noha Sharafeldin, Ofer Sadan, Patricia A. Francis, Penny Wung Burgoon, Peter Robinson, Philip R.O. Payne, Rafael Fuentes, Randeep Jawa, Rebecca Erwin-Cohen, Rena Patel, Richard A. Moffitt, Richard L. Zhu, Rishi Kamaleswaran, Robert Hurley, Robert T. Miller, Saiju Pyarajan, Sam G. Michael, Samuel Bozzette, Sandeep Mallipattu, Satyanarayana Vedula, Scott Chapman, Shawn T. O’Neil, Soko Setoguchi, Stephanie S. Hong, Steve Johnson, Tellen D. Bennett, Tiffany Callahan, Umit Topaloglu, Usman Sheikh, Valery Gordon, Vignesh Subbian, Warren A. Kibbe, Wenndy Hernandez, Will Beasley, Will Cooper, William Hillegass, Xiaohan Tanner Zhang. Details of contributions available at covid.cd2h.org/core-contributors

## Data Partners with Released Data

The following institutions whose data is released or pending: Available: Advocate Health Care Network — UL1TR002389: The Institute for Translational Medicine (ITM) • Boston University Medical Campus — UL1TR001430: Boston University Clinical and Translational Science Institute • Brown University — U54GM115677: Advance Clinical Translational Research (Advance-CTR) • Carilion Clinic — UL1TR003015: iTHRIV Integrated Translational health Research Institute of Virginia • Charleston Area Medical Center — U54GM104942: West Virginia Clinical and Translational Science Institute (WVCTSI) • Children’s Hospital Colorado — UL1TR002535: Colorado Clinical and Translational Sciences Institute • Columbia University Irving Medical Center — UL1TR001873: Irving Institute for Clinical and Translational Research • Duke University — UL1TR002553: Duke Clinical and Translational Science Institute • George Washington Children’s Research Institute — UL1TR001876: Clinical and Translational Science Institute at Children’s National (CTSA-CN) • George Washington University — UL1TR001876: Clinical and Translational Science Institute at Children’s National (CTSA-CN) • Indiana University School of Medicine — UL1TR002529: Indiana Clinical and Translational Science Institute • Johns Hopkins University — UL1TR003098: Johns Hopkins Institute for Clinical and Translational Research • Loyola Medicine — Loyola University Medical Center • Loyola University Medical Center — UL1TR002389: The Institute for Translational Medicine (ITM) • Maine Medical Center — U54GM115516: Northern New England Clinical & Translational Research (NNE-CTR) Network • Massachusetts General Brigham — UL1TR002541: Harvard Catalyst • Mayo Clinic Rochester — UL1TR002377: Mayo Clinic Center for Clinical and Translational Science (CCaTS) • Medical University of South Carolina — UL1TR001450: South Carolina Clinical & Translational Research Institute (SCTR) • Montefiore Medical Center — UL1TR002556: Institute for Clinical and Translational Research at Einstein and Montefiore • Nemours U54GM104941: Delaware CTR ACCEL Program • NorthShore University HealthSystem — UL1TR002389: The Institute for Translational Medicine (ITM) • Northwestern University at Chicago — UL1TR001422: Northwestern University Clinical and Translational Science Institute (NUCATS) • OCHIN — INV-018455: Bill and Melinda Gates Foundation grant to Sage Bionetworks • Oregon Health & Science University — UL1TR002369: Oregon Clinical and Translational Research Institute • Penn State Health Milton S. Hershey Medical Center — UL1TR002014: Penn State Clinical and Translational Science Institute • Rush University Medical Center — UL1TR002389: The Institute for Translational Medicine (ITM) • Rutgers, The State University of New Jersey — UL1TR003017: New Jersey Alliance for Clinical and Translational Science • Stony Brook University — U24TR002306 • The Ohio State University — UL1TR002733: Center for Clinical and Translational Science • The State University of New York at Buffalo — UL1TR001412: Clinical and Translational Science Institute • The University of Chicago — UL1TR002389: The Institute for Translational Medicine (ITM) • The University of Iowa — UL1TR002537: Institute for Clinical and Translational Science • The University of Miami Leonard M. Miller School of Medicine — UL1TR002736: University of Miami Clinical and Translational Science Institute • The University of Michigan at Ann Arbor — UL1TR002240: Michigan Institute for Clinical and Health Research • The University of Texas Health Science Center at Houston — UL1TR003167: Center for Clinical and Translational Sciences (CCTS) • The University of Texas Medical Branch at Galveston — UL1TR001439: The Institute for Translational Sciences • The University of Utah — UL1TR002538: Uhealth Center for Clinical and Translational Science • Tufts Medical Center — UL1TR002544: Tufts Clinical and Translational Science Institute • Tulane University — UL1TR003096: Center for Clinical and Translational Science • University Medical Center New Orleans — U54GM104940: Louisiana Clinical and Translational Science (LA CaTS) Center • University of Alabama at Birmingham — UL1TR003096: Center for Clinical and Translational Science • University of Arkansas for Medical Sciences — UL1TR003107: UAMS Translational Research Institute • University of Cincinnati — UL1TR001425: Center for Clinical and Translational Science and Training • University of Colorado Denver, Anschutz Medical Campus — UL1TR002535: Colorado Clinical and Translational Sciences Institute • University of Illinois at Chicago — UL1TR002003: UIC Center for Clinical and Translational Science • University of Kansas Medical Center — UL1TR002366: Frontiers: University of Kansas Clinical and Translational Science Institute • University of Kentucky — UL1TR001998: UK Center for Clinical and Translational Science • University of Massachusetts Medical School Worcester — UL1TR001453: The UMass Center for Clinical and Translational Science (UMCCTS) • University of Minnesota — UL1TR002494: Clinical and Translational Science Institute • University of Mississippi Medical Center — U54GM115428: Mississippi Center for Clinical and Translational Research (CCTR) • University of Nebraska Medical Center — U54GM115458: Great Plains IDeA-Clinical & Translational Research • University of North Carolina at Chapel Hill — UL1TR002489: North Carolina Translational and Clinical Science Institute • University of Oklahoma Health Sciences Center — U54GM104938: Oklahoma Clinical and Translational Science Institute (OCTSI) • University of Rochester — UL1TR002001: UR Clinical & Translational Science Institute • University of Southern California — UL1TR001855: The Southern California Clinical and Translational Science Institute (SC CTSI) • University of Vermont — U54GM115516: Northern New England Clinical & Translational Research (NNE-CTR) Network • University of Virginia — UL1TR003015: iTHRIV Integrated Translational health Research Institute of Virginia • University of Washington — UL1TR002319: Institute of Translational Health Sciences • University of Wisconsin-Madison — UL1TR002373: UW Institute for Clinical and Translational Research • Vanderbilt University Medical Center — UL1TR002243: Vanderbilt Institute for Clinical and Translational Research • Virginia Commonwealth University — UL1TR002649: C. Kenneth and Dianne Wright Center for Clinical and Translational Research • Wake Forest University Health Sciences — UL1TR001420: Wake Forest Clinical and Translational Science Institute • Washington University in St. Louis — UL1TR002345: Institute of Clinical and Translational Sciences • Weill Medical College of Cornell University — UL1TR002384: Weill Cornell Medicine Clinical and Translational Science Center • West Virginia University — U54GM104942: West Virginia Clinical and Translational Science Institute (WVCTSI) Submitted: Icahn School of Medicine at Mount Sinai — UL1TR001433: ConduITS Institute for Translational Sciences • The University of Texas Health Science Center at Tyler — UL1TR003167: Center for Clinical and Translational Sciences (CCTS) • University of California, Davis — UL1TR001860: UCDavis Health Clinical and Translational Science Center • University of California, Irvine — UL1TR001414: The UC Irvine Institute for Clinical and Translational Science (ICTS) • University of California, Los Angeles — UL1TR001881: UCLA Clinical Translational Science Institute • University of California, San Diego — UL1TR001442: Altman Clinical and Translational Research Institute • University of California, San Francisco — UL1TR001872: UCSF Clinical and Translational Science Institute Pending: Arkansas Children’s Hospital — UL1TR003107: UAMS Translational Research Institute • Baylor College of Medicine — None (Voluntary) • Children’s Hospital of Philadelphia — UL1TR001878: Institute for Translational Medicine and Therapeutics • Cincinnati Children’s Hospital Medical Center — UL1TR001425: Center for Clinical and Translational Science and Training • Emory University — UL1TR002378: Georgia Clinical and Translational Science Alliance • HonorHealth — None (Voluntary) • Loyola University Chicago — UL1TR002389: The Institute for Translational Medicine (ITM) • Medical College of Wisconsin — UL1TR001436: Clinical and Translational Science Institute of Southeast Wisconsin • MedStar Health Research Institute — UL1TR001409: The Georgetown-Howard Universities Center for Clinical and Translational Science (GHUCCTS) • MetroHealth — None (Voluntary) • Montana State University — U54GM115371: American Indian/Alaska Native CTR • NYU Langone Medical Center — UL1TR001445: Langone Health’s Clinical and Translational Science Institute • Ochsner Medical Center — U54GM104940: Louisiana Clinical and Translational Science (LA CaTS) Center • Regenstrief Institute — UL1TR002529: Indiana Clinical and Translational Science Institute • Sanford Research — None (Voluntary) • Stanford University — UL1TR003142: Spectrum: The Stanford Center for Clinical and Translational Research and Education • The Rockefeller University — UL1TR001866: Center for Clinical and Translational Science • The Scripps Research Institute — UL1TR002550: Scripps Research Translational Institute • University of Florida — UL1TR001427: UF Clinical and Translational Science Institute • University of New Mexico Health Sciences Center — UL1TR001449: University of New Mexico Clinical and Translational Science Center • University of Texas Health Science Center at San Antonio — UL1TR002645: Institute for Integration of Medicine and Science • Yale New Haven Hospital — UL1TR001863: Yale Center for Clinical Investigation

## Authors statement

Authorship was determined using ICMJE recommendations.

ZB: Generated research question, drafted manuscript, managed project timeline, and coordinated analysis.

YJ, SD, EH, JC, and JS: Reviewed manuscript, provided feedback, and supported analysis.

AM, ML, JC, RP, and AH: Provided oversight on study design and analysis plan, reviewed manuscript, provided feedback, and supported interpretations.

