## Appendix 1 for "SSRI Use During Acute COVID-19 Infection Associated with Lower Risk of Long COVID Among Patients with Depression"

**Appendix 1. Covariate information.**

Unless otherwise stated, all variables were evaluated before baseline (index acute COVID-19 infection).

Selective serotonin reuptake inhibitor (SSRI) use (code sets 602742506, 709408353, 402078260, 738475452, 255742597, 408043043, 444624587): We defined SSRI users as individuals who were using an SSRI medication (fluoxetine, sertraline, paroxetine, citalopram, escitalopram, fluvoxamine, and vilazodone, phenotyped using RxNorm) at the time of COVID-19 infection (beginning at least 30 days prior to COVID-19 infection) and we defined all other individuals as nonusers.

Long COVID Diagnosis (post-acute sequelae of COVID-19, PASC, ICD-10 code U09.9): We defined Long COVID diagnosis as between 1 and 12 months following acute COVID-19 infection.

Number of visits pre-COVID-19 infection: The number of healthcare visits that a participant attended between the start of N3C monitoring (January 1, 2018) and incident COVID-19 infection. We define a healthcare visit as an interaction (or cluster of interactions) with a healthcare provider that was associated with a given medical condition, diagnosis, or procedure.

Healthcare visits per month before COVID-19 infection): The number of healthcare visits that a participant attended between the start of N3C monitoring (January 1, 2018) and incident COVID-19 infection divided by the number of months between their first medical visit documented in N3C and their incident COVID-19 infection.

Sex: Participant biologic sex.

Age at acute COVID-19 infection: Patient age in years.

Race/ ethnicity: A covariate that combines a patient’s categorical race status and categorical ethnicity.

Common data model format: The common data model documentation format used by the contributing data provider.

Region of residence: A categorical measure of geographic region, denoted by the first digits of the patient zip code.

Body mass index (BMI): A continuous measure of body composition.

Tobacco smoking status: A binary indicator denoting current tobacco smoking status (user vs. nonuser)

Obesity: A binary indicator denoting a previous diagnosis of obesity.

Diabetes: A binary indicator denoting a previous diagnosis of diabetes.

Chronic lung disease: A binary indicator denoting a previous diagnosis of chronic lung disease.

Heart failure: A binary indicator denoting a previous diagnosis with heart failure.

Hypertension: A binary indicator of hypertension diagnosis.

Use of systemic corticosteroids: A binary indicator of whether a patient was currently receiving systemic corticosteroids.

Whether the patient was immunocompromised: A binary indicator variable denoting whether the patient was diagnosed with having a comprised immune status.

Number of COVID-19 vaccination doses before infection: The number of COVID-19 vaccination and booster doses that the patient received.

Percent of the county with an income level below the poverty line: County-level variable indicating what percent of the county received an income below the federal poverty line.

Social deprivation index score: County-level variable that quantifies social deprivation and inequality.

Depression severity score (code sets 649312011, 924814934, 937942059): A three-level categorical variable denoting the patient’s severity of major depressive disorder. Levels include “mild,” “moderate,” and “severe.” Depression severity was evaluated prior to SSRI use.

Bipolar disorder: An indicator variable denoting patient diagnosis with bipolar disorder.

Post-COVID visit indicator: An indicator variable denoting whether the patient had a documented healthcare visit between 1 and 12 months following index acute COVID-19 infection (the outcome observation period).

Bone fracture (code set 889591431): A binary variable indicating a bone fracture between 1 and 12 months following COVID-19 infection.
